# Changes in respiratory infection trends during the COVID-19 pandemic in the haematologic malignancy patients

**DOI:** 10.1101/2023.08.29.23294751

**Authors:** Jiwon RYOO, Seok Chan Kim, Jongmin Lee

## Abstract

**Background:** The coronavirus disease 2019 (COVID-19) pandemic globally changed respiratory infection patterns. However, its impact on community-acquired pneumonia (CAP) in high risk patients with haematological malignancies (HM) is uncertain. We aimed to examine CAP aetiology changes in patients with HM pre- and post-COVID-19 pandemic.

**Methods:** This retrospective study included 524 HM patients hospitalised with CAP between March 2018 and February 2022. Those who underwent bronchoscopy within 24 hours after admission to identify CAP aetiology were included. Data on patient characteristics, laboratory findings, and results of bronchioalveolar lavage fluid cultures and PCR tests were analysed to compare etiological changes and identify in-hospital mortality risk factors.

**Results:** Patients were divided into pre-COVID-19 (44.5%) and post-COVID-19 (55.5%) groups. This study found a significant decrease in viral CAP in the post-COVID-19 era, particularly for influenza A, parainfluenza, adenovirus, and rhinovirus (3.0% vs. 0.3%, respectively, P = 0.036; 6.5% vs. 0.7%, respectively, P = 0.001; 5.6% vs. 1.4%, respectively, P = 0.015; 9.5% vs. 1.7%, respectively, P < 0.001). Bacterial, fungal, and unknown CAP aetiologies remain unchanged. Higher Sequential Organ Failure Assessment scores and lower platelet count correlated with in-hospital mortality after adjusting for potential confounding factors.

**Conclusion:** The incidence of CAP in HM patients did not decrease after COVID-19. Additionally, CAP aetiology among patients with HM changed following the COVID-19 pandemic, with a significant reduction in viral pneumonia while bacterial and fungal pneumonia persisted. Further studies are required to evaluate the impact of COVID-19 on the prognosis of patients with HM and CAP.

## Background

The coronavirus disease 2019 (COVID-19) pandemic led to significant changes in respiratory infection patterns. After its outbreak in December 2019, in Wuhan, China, COVID-19 spread rapidly worldwide. Governments globally have extensively advocated for a range of measures to curb the COVID-19 pandemic, such as wearing masks, education on hand hygiene, social distancing, and travel restrictions. The Korean government has implemented several measures since February 2020 to prevent the COVID-19 outbreak, including active epidemiological investigations, quarantine of suspicious cases, and extensive public lockdowns[1]. The implementation of these strategies resulted in not only a decrease in the spread of COVID-19 but also substantially reduced other respiratory infections. Previous studies have reported a reduction in seasonal influenza activity [2-4], leading to a subsequent decrease in the occurrence of community-acquired pneumonia and hospital as well as ICU (intensive care unit) admissions [5].

Owing to both the characteristics of their condition and antineoplastic therapies, patients with haematological malignancies (HM) are immunocompromised and at high risk of infectious complications. They have a high incidence of community-acquired pneumonia (CAP), leading to substantial morbidity and mortality within this population [6,7]. Early identification of CAP aetiology in patients with HM is important; however, despite the use of a comprehensive diagnostic workup, the aetiology of pneumonia is often under identified [8,9]. Previous studies have revealed that immunocompromised patients with pneumonia of an undetermined aetiology have increased mortality rates [10,11]. Furthermore, due to their immunocompromised status, the aetiology of CAP in these patients presents distinct characteristics compared to that in the general population. Therefore, a thorough exploration of the changing patterns in the aetiology of CAP, specifically in patients with HM, is warranted. Moreover, the impact of the COVID-19 era on the changing patterns of CAP in patients with HM remains uncertain. Therefore, this study aimed to examine the changes in CAP aetiology in patients with HM before and after the COVID-19 pandemic.

## Methods

### Patients

We retrospectively studied a cohort of patients with HM who visited the emergency department or outpatient clinic of the Seoul St. Mary’s Hospital (Seoul, Korea) with respiratory symptoms and required admission between March 2018 and February 2022. In this hospital, more than 500 patients undergo haematopoietic stem cell transplantations (HSCTs) annually. Eligible patients included those who exhibited abnormal findings on chest X-ray and underwent bronchoscopy (BRS) within 24 hours after admission. Patients who did not undergo BRS within 24 hours after admission, were diagnosed with hospital-acquired pneumonia (HAP), or those newly diagnosed with COVID-19 were excluded (Figure 1). Patients with HM were defined as those who underwent concurrent evaluations and treatment without remission [12]. This study was approved by the Institutional Review Board of Seoul St. Mary’s Hospital (KC23RISI0113), and the requirement for informed consent was waived.

**Figure 1.**
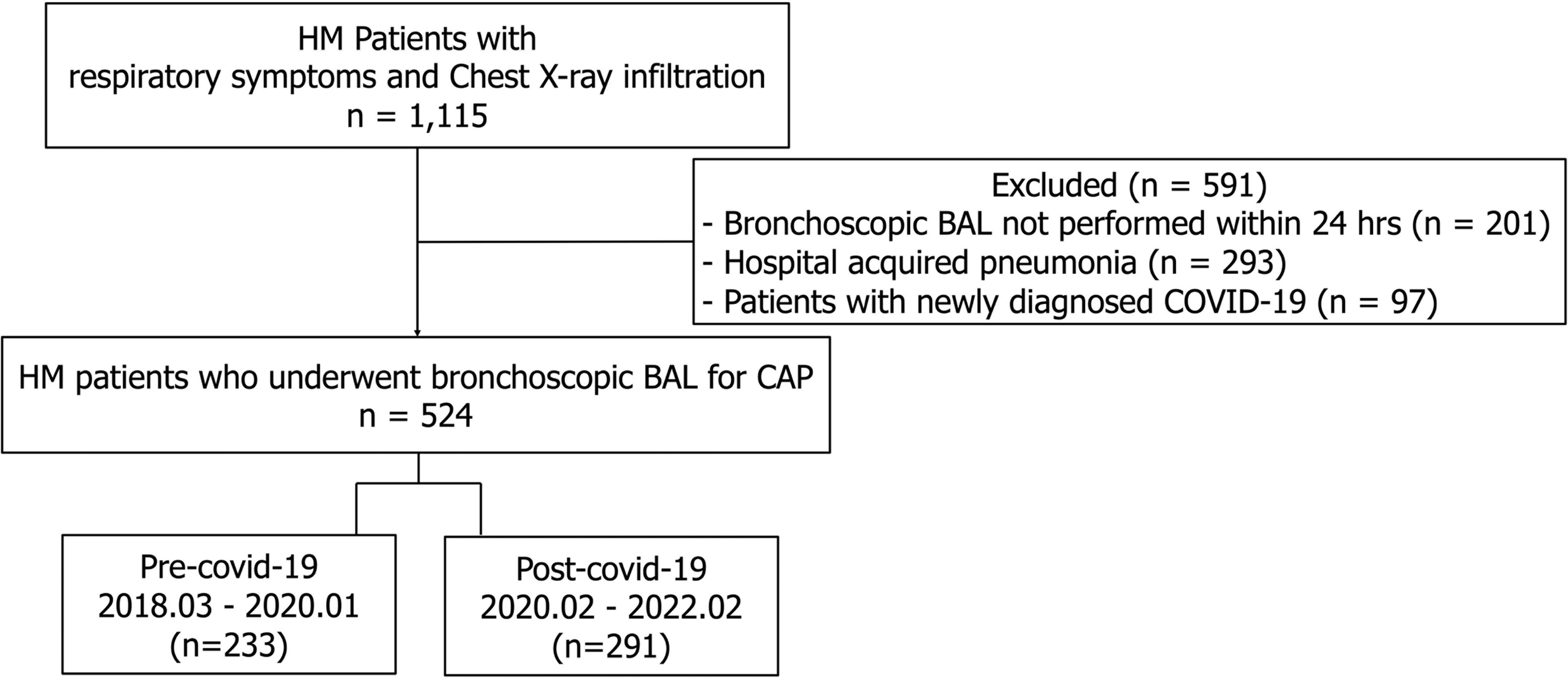
Study flow diagram. HM, haematologic malignancy; BAL, bronchoalveolar lavage; COVID-19, coronavirus disease 2019; CAP, community-acquired pneumonia.

### Data collection

Epidemiological and clinical data were collected from the patients’ medical charts at admission. Data included sex; age; laboratory findings, including white blood cell counts (WBC), absolute neutrophil counts (ANC), absolute lymphocyte counts (ALC), haemoglobin, haematocrit, platelet, and C-reactive protein (CRP) levels; characteristics of the haematologic malignancies, including the type of malignancy, current status, and prior treatments; Sequential Organ Failure Assessment (SOFA) score at admission; body temperature at admission; and pattern of chest X-ray abnormalities. The results of sputum cultures from bronchoalveolar lavage (BAL) fluid culture, BAL galactomannan test, special staining (Gomori methenamine silver, periodic acid Schiff, or Ziehl-Neelsen) of BAL cells, atypical pneumonia serological panel (*Chlamydia, Legionella,* and *Mycoplasma*), blood culture, and serum galactomannan were also reviewed. A respiratory virus polymerase chain reaction (RV PCR) multiplex panel (AdvanSure RV real-time PCR Kit; LG Life Sciences, Seoul, Korea) was used to test for influenza A and B viruses, parainfluenza (PIV), respiratory syncytial virus (RSV), adenovirus (ADV), bocavirus, human metapneumovirus (MPV), coronavirus, and human rhinovirus (HRV) for BAL samples. Nucleic acid extraction was performed using a QIAamp DNA Mini Kit in an automated extractor (Qiacube, Qiagen, Hilden, Germany).

Pneumonia was defined as the presence of a new infiltration on a chest radiograph at the time of the hospital visit with more than one of the following criteria: (1) new or increased cough with or without sputum production, (2) fever (temperature ≥38.0_J) or hypothermia (<35.0_J), and (3) evidence of systemic inflammation (abnormal white blood cell count or increased CRP) [6]. Invasive fungal diseases, including pulmonary aspergillosis (IPA), were defined by the presence of compatible host factors, clinical features, and mycological evidence according to the European Organisation for Research and Treatment of Cancer/Mycoses Study Group criteria [13]. The diagnosis of *Pneumocystis jirovecii* pneumonia was based on the identification of the organism and/or PCR in the BAL fluid, along with compatible clinical features and radiological findings [13]. CAP cases of unknown aetiology were characterised by negative results in both BAL culture and PCR tests as well as negative serological examinations where no other identifiable cause of pulmonary infiltration could be found. Patients with HAP were defined as those admitted to the hospital for a reason other than acute respiratory infection, in whom respiratory symptoms developed ≥72 hours after admission. Finally, we examined the clinical outcomes, including ICU admission and in-hospital mortality.

### Statistical analysis

Continuous variables were reported as median (range), whereas categorical variables were described as numbers (%). Patient characteristics were compared using the chi-squared test or Fisher’s exact test, as appropriate, for categorical variables and independent sample t-tests for continuous variables. The odds ratios (OR) and their corresponding intervals (CI) were computed. Goodness-of-fit was computed to assess the relevance of the logistic regression model. All tests were two-sided, and a P value <0.05 was considered statistically significant. All statistical analyses were performed using SPSS for Windows software (ver. 20.0; IBM Corp., Armonk, NY, USA) and R (ver. 4.3.1, R Foundation, Venna, Austria).

### Patient and public involvement

Due to the retrospective design of this study, neither patients nor the public were involved. However, we anticipate that our findings will benefit future patient cohorts.

## Results

### Patient characteristics

Among the 1,115 patients with HM, respiratory symptoms, and chest X-ray infiltration admitted to our hospital between March 2018 and February 2022, 524 were included in our analysis (Figure 1). As Korea implemented COVID-19 containment policies in February 2020, we divided the timeline into pre-COVID-19 and post-COVID-19 eras, using February 2020 as the point of demarcation [1]. pre-COVID-19 and post-COVID-19 groups comprised 233 (44.5%) and 291 (55.5%) patients, respectively. Aside from a higher proportion of patients with bilateral pulmonary infiltration in their chest X-rays within the pre-COVID-19 group (81.1% vs. 69.4%, respectively, P = 0.003), no significant differences were observed between the two groups in terms of patient characteristics (Table 1).

**Table 1.**
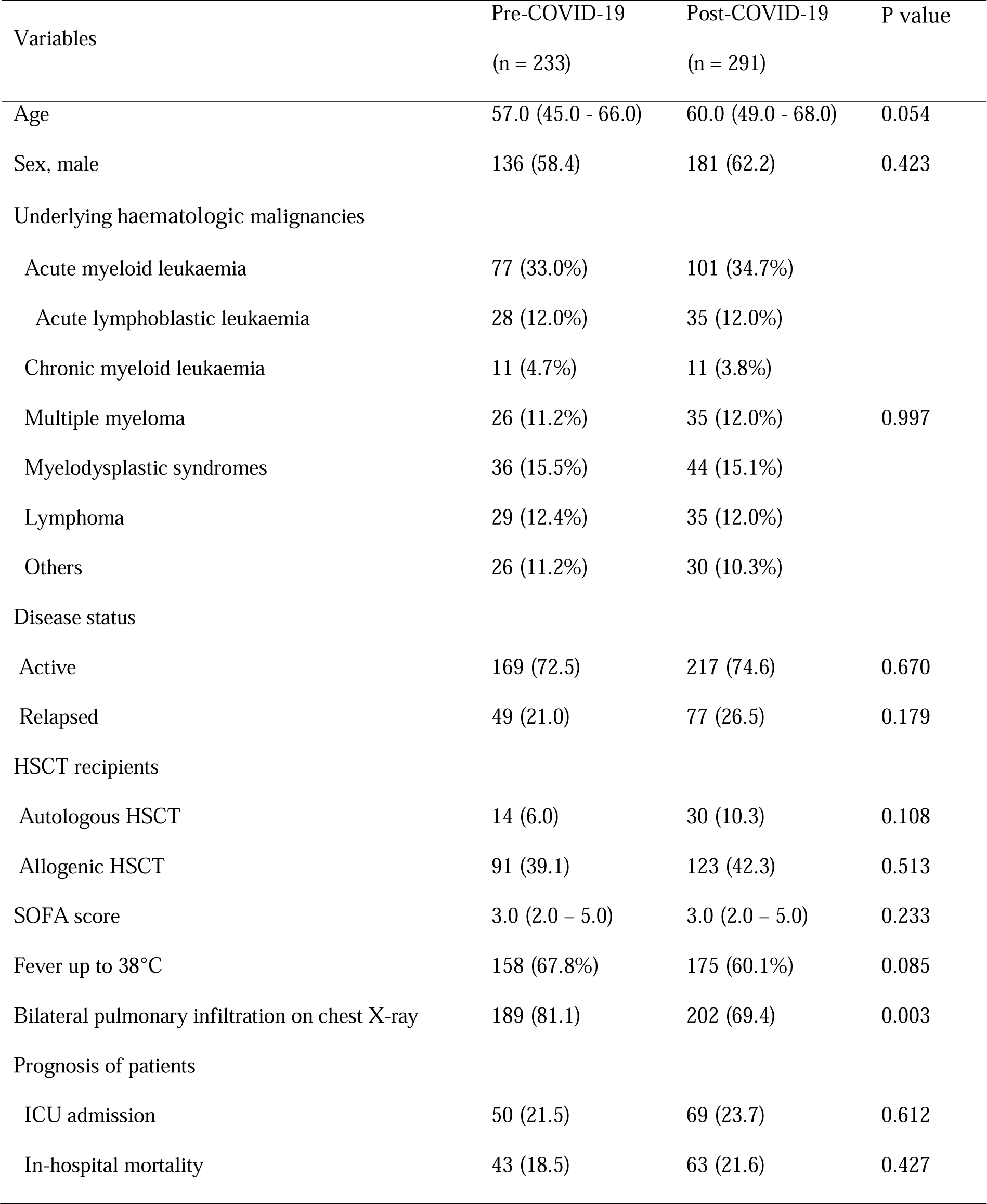
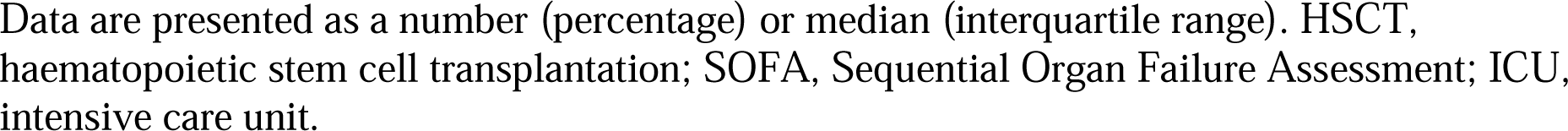
Baseline characteristics.

### Changes in the aetiology of community-acquired pneumonia in patients with haematologic malignancies

As of February 2020, no significant differences were evident in the number of patients admitted due to CAP when dividing the population into pre-COVID-19 and post-COVID-19 groups (Figure 2). Table 2 and Figure 3 show the changes in the aetiology of CAP after the pandemic. Prior to it, the proportions of aetiologies were as follows: unknown aetiology (36.5%), respiratory virus (30.7%), bacteria (23.2%), and fungus (16.3%). During the post-COVID-19 era, the proportion of aetiologies changed as follows: unknown aetiology (41.6%), bacteria (24.4%), fungus (18.9%), and respiratory virus (6.6%). In the pre- and post-COVID-19 pandemic periods, the proportions of bacterial, fungal, and unknown aetiology CAP remained unchanged, whereas viral CAP significantly decreased. As shown in Table 2 and Figure 4, the proportion of respiratory viruses decreased after the pandemic. In particular, a significant reduction in the incidence of influenza A (3.0% vs. 0.3%, respectively, P = 0.036), parainfluenza (6.5% vs. 0.7%, respectively, P = 0.001), adenovirus (5.6% vs. 1.4%, respectively, P = 0.015), and rhinovirus (9.5% vs. 1.7%, respectively, P < 0.001) were observed after the pandemic.

**Figure 2.**
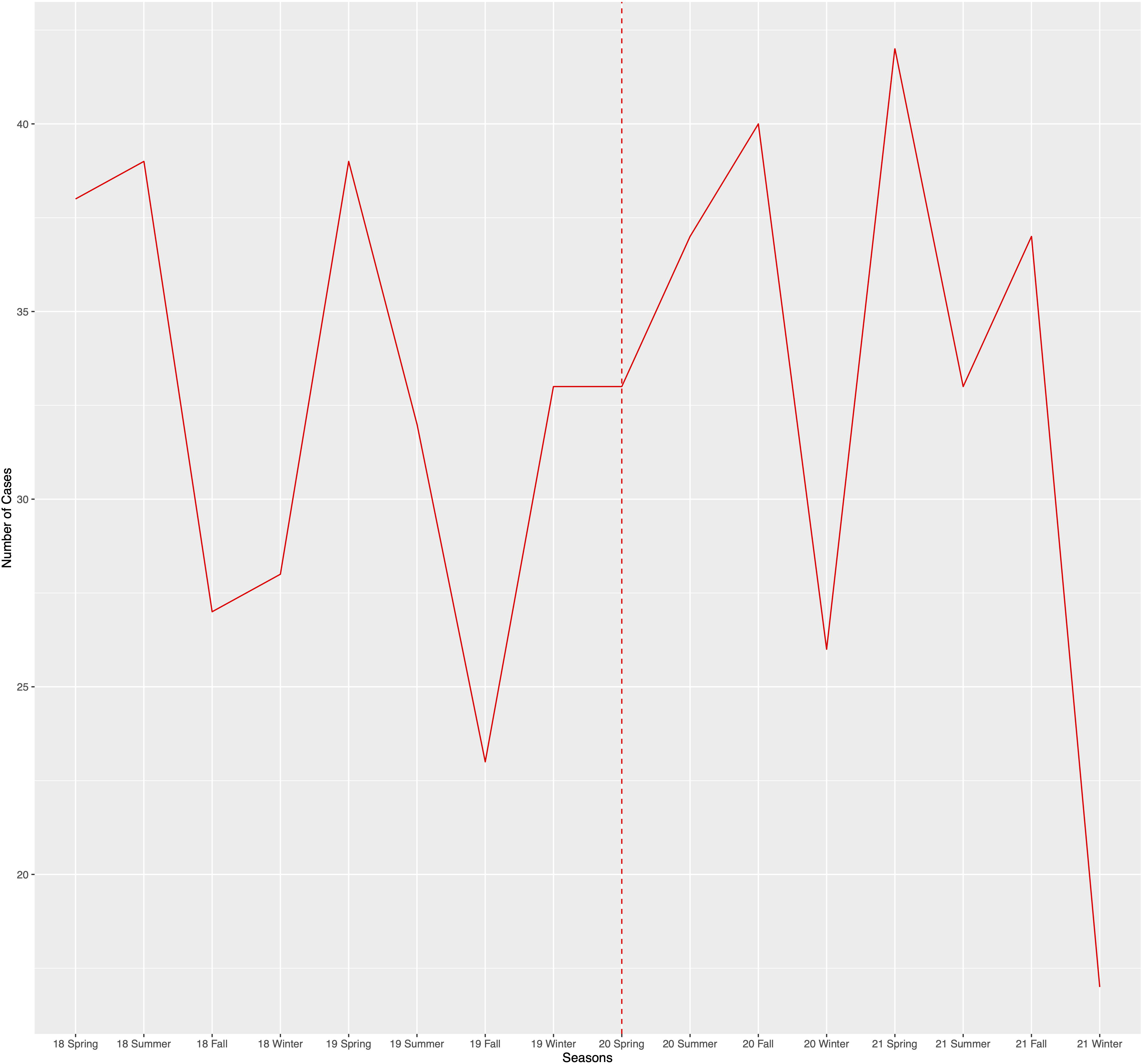
The total number of admitted patients with CAP between March 2019 and February 2022. Spring, from March to May; Summer, from June to August; Fall, from September to November; Winter, from December to February.

**Figure 3.**
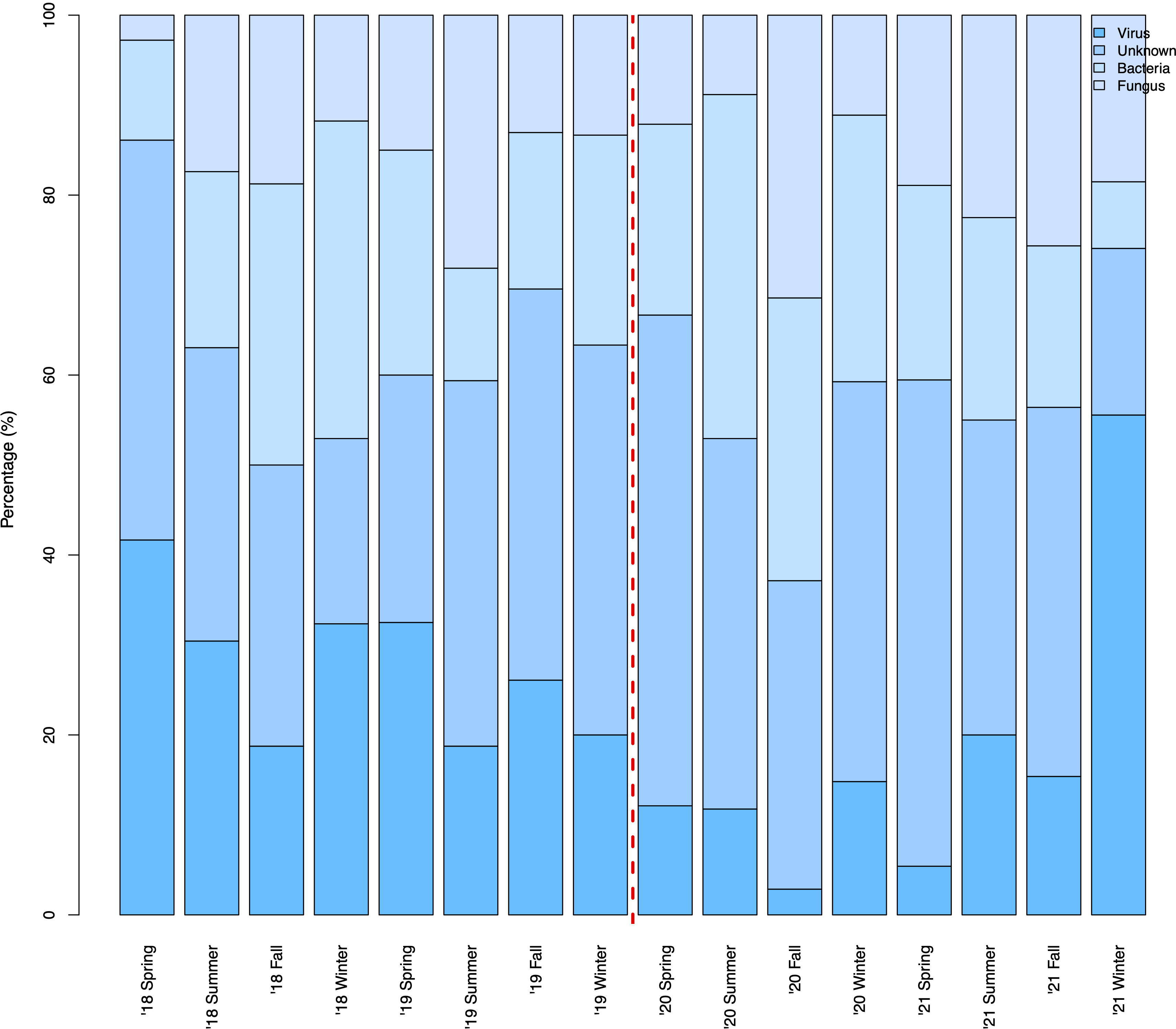
The change in CAP aetiology over time. Spring, from March to May; Summer, from June to August; Fall, from September to November; Winter, from December to February.

**Figure 4.**
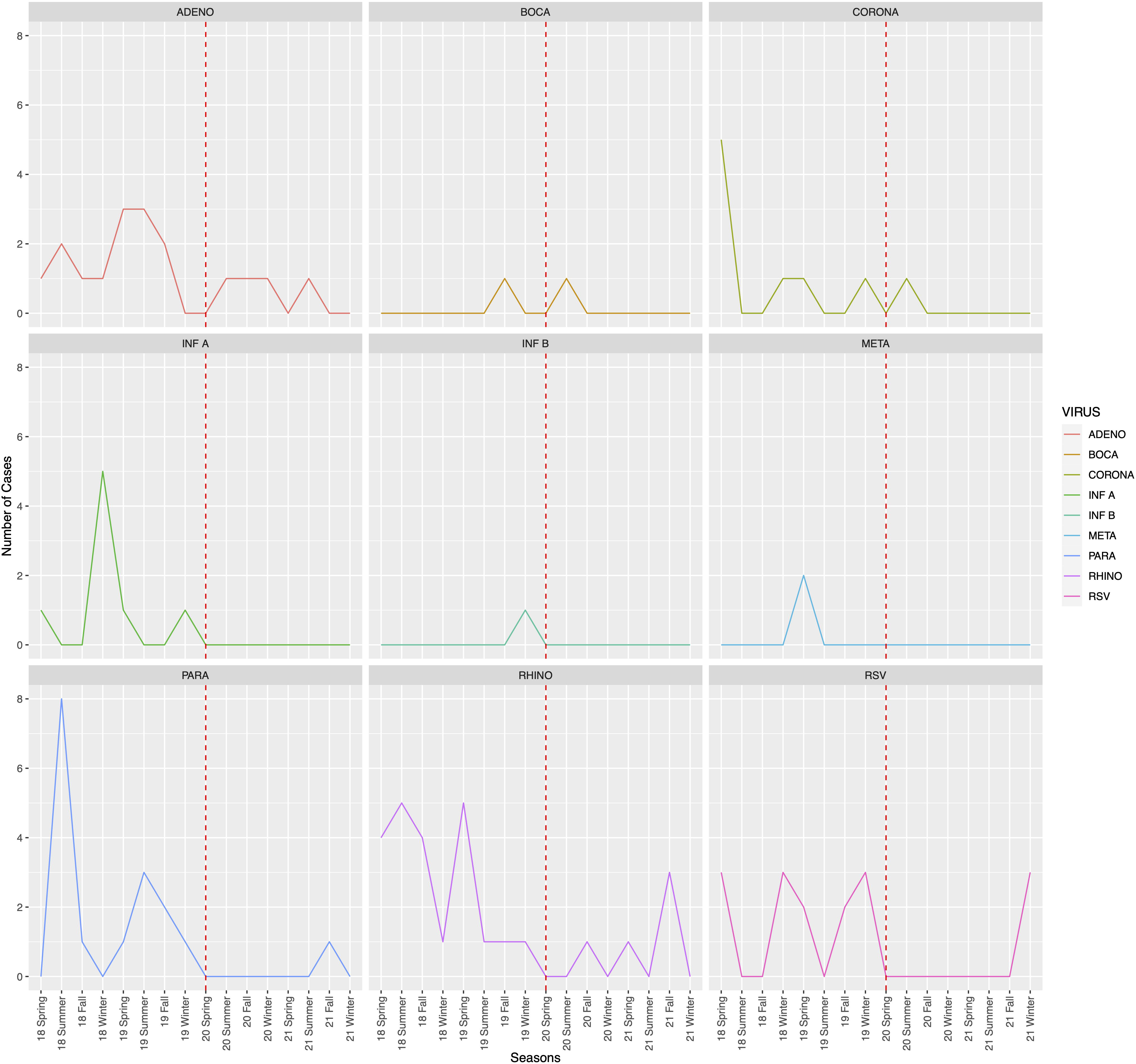
Seasonal distribution of respiratory virus detection cases. ADENO, adenovirus; BOCA, bocavirus; CORONA, human coronavirus; INF A, Influenza A; INF B, Influenza B; META, metapneumovirus; PARA, parainfluenza virus; RHINO, rhinovirus; RSV, respiratory syncytial virus. Spring, from March to May; Summer, from June to August; Fall, from September to November; Winter, from December to February.

**Table 2.**
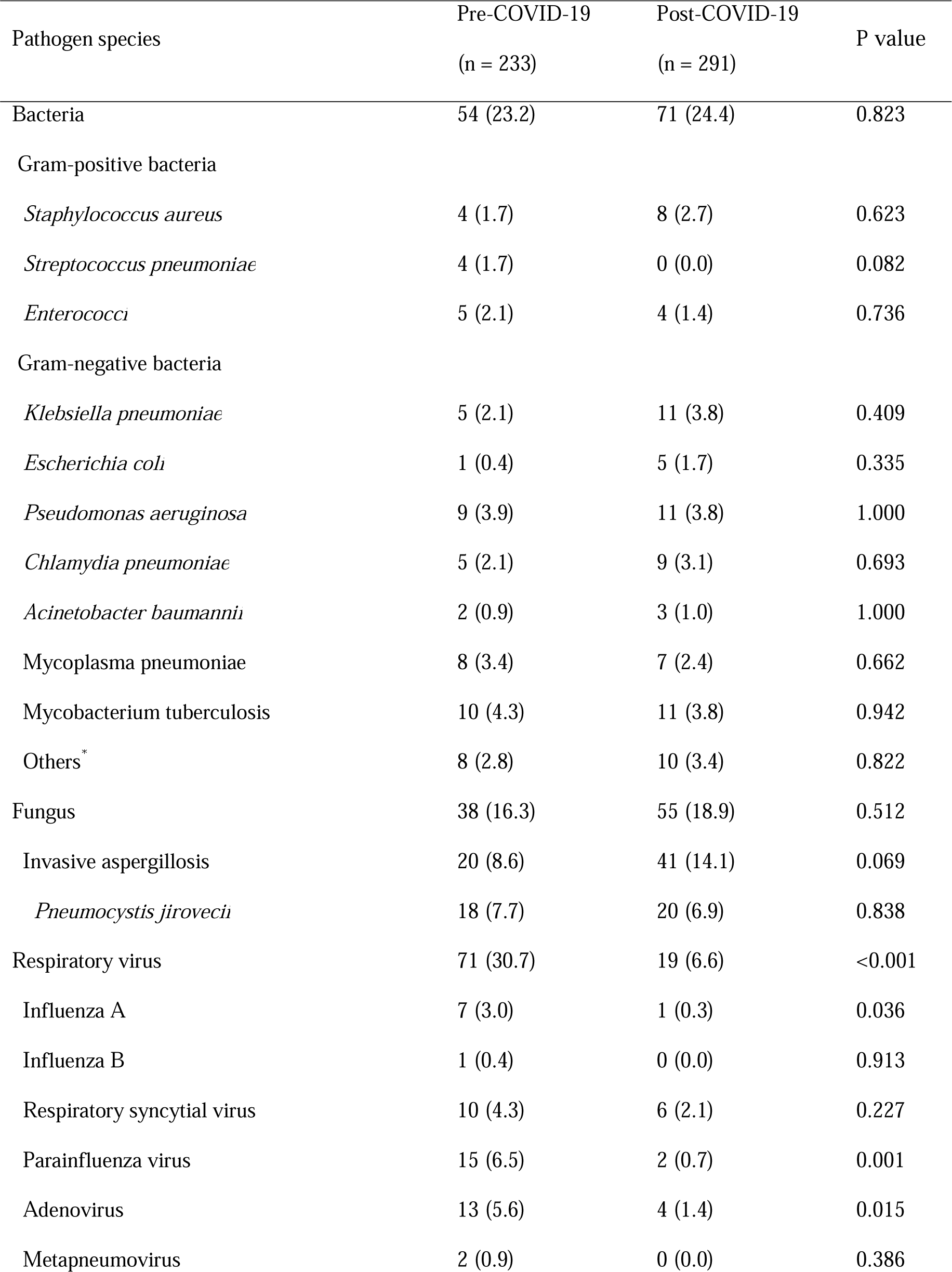
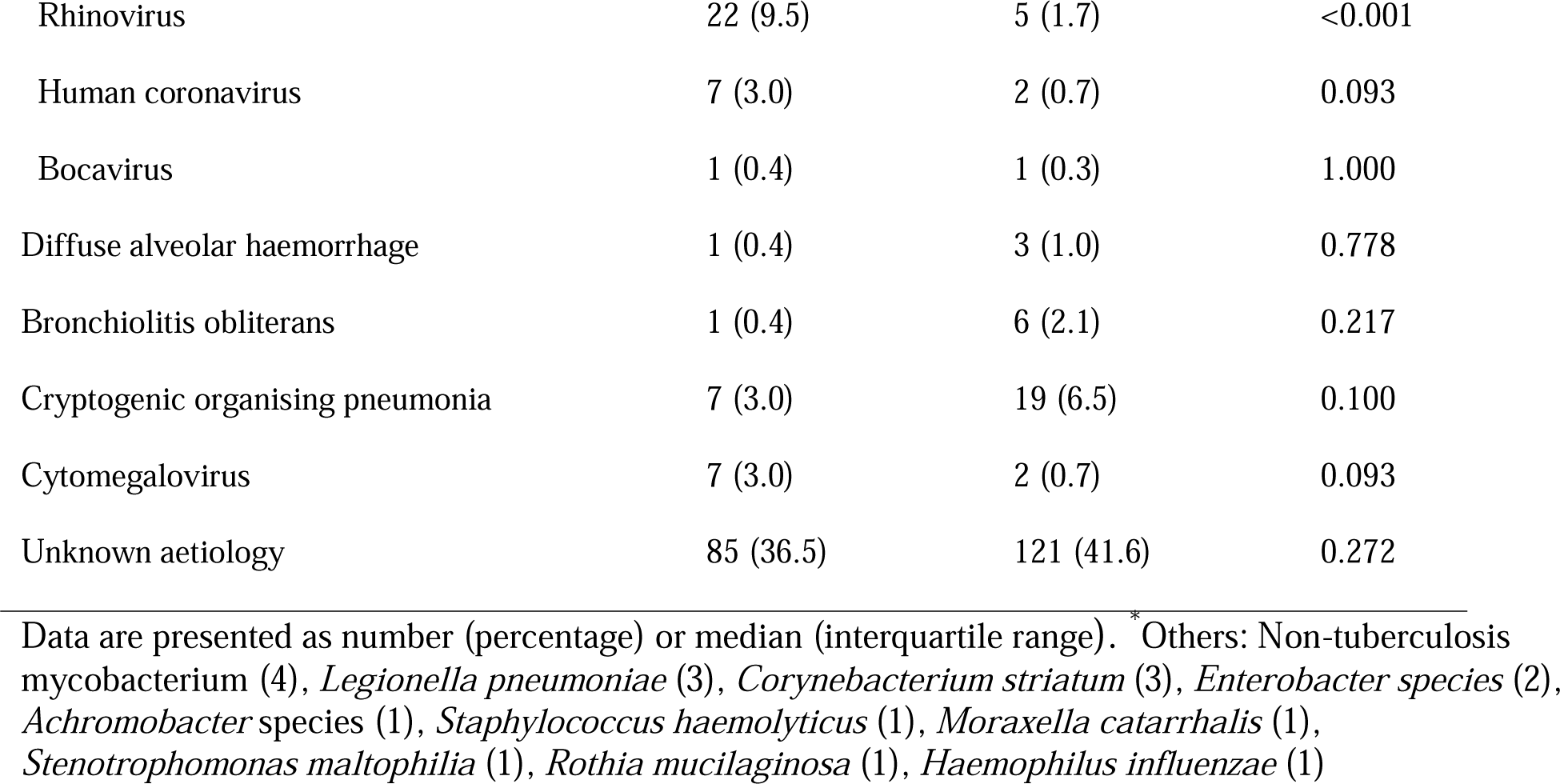
The Differential Aetiology of Pneumonia Pre- and Post-COVID-19 Outbreak.

### Factors associated with increased mortality

The S1 Table presents comparisons of the clinical characteristics between in-hospital deaths and survivors. Among non-survivors, a higher proportion of patients with active HM diseases (83.0% vs. 71.3%, respectively, P = 0.020), CRP (12.2 [6.4 – 21.3] vs. 8.4 [3.7 – 17.5], respectively, P = 0.001), relapsed diseases (34.9% vs. 21.3%, respectively, P = 0.005), and SOFA scores (4.0 [2.0–5.0] vs. 3.0 [2.0–5.0], respectively, P < 0.001), and lower ANCs (1.4 [0.2–6.1] vs. 3.3 [0.6–6.4], respectively, P = 0.049), haemoglobin (8.6 [7.6-10,60] vs. 9.7 [8.4–11.8], respectively, P < 0.001), haematocrit (25.1 [22.1–31.2] vs. 29.3 [25.2–35.5], respectively, P < 0.001), and lower platelet count (27.5 [14.0–73.0] vs. 98.0 [36.0 – 191.0], respectively, P < 0.001) were found.

The logistic regression analysis of the clinical parameters used to evaluate the risk factors associated with in-hospital mortality is shown in Table 3. A high SOFA score and CRP level, an active disease, and a low haemoglobin level, haematocrit level, and platelet count were independent risk factors for hospital mortality. After adjusting for potential confounding factors, higher SOFA scores and lower platelet counts were independently associated with in-hospital mortality (P < 0.001 and P = 0.004, respectively).

**Table 3.**
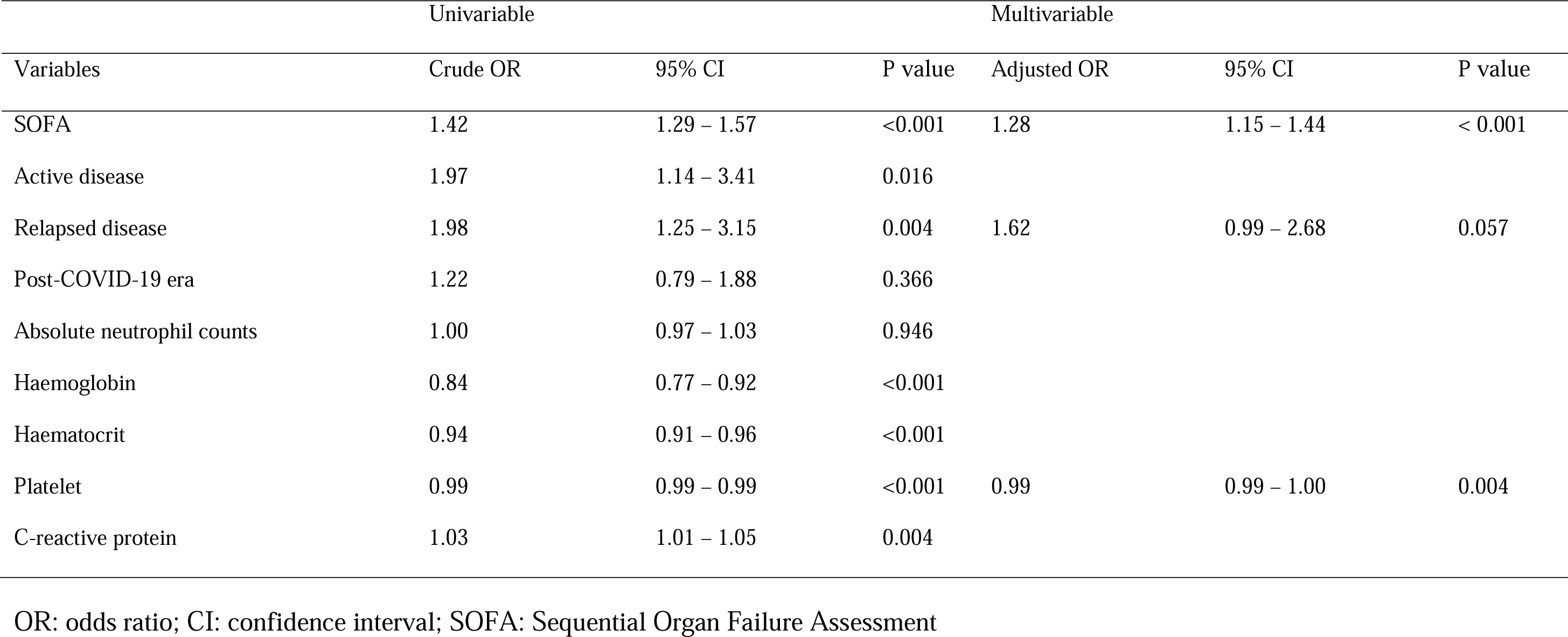
Logistic regression analysis for in-hospital mortality.

## Discussion

In the present study, we compared changes in the aetiology of CAP among patients with HM before and after the COVID-19 pandemic. After the pandemic, numerous studies have focused on the incidence and treatment of COVID-19 in immunocompromised patients; however, limited research has been conducted on the changes in aetiology other than COVID-19 that can cause CAP in such patient populations. Nonetheless, considering the susceptibility of immunocompromised patients to various opportunistic infections, it is crucial to examine the changes in pathogens that can cause CAP after COVID-19.

In contrast to the findings of previous studies that reported a decrease in the occurrence of CAP after COVID-19 [14-17], in this study, the number of HM patients hospitalised with CAP during the post-COVID-19 era did not decrease compared to the that during the pre-COVID-19 era. Notably, viral pneumonia demonstrated a significant decrease; however, bacterial and fungal pneumonia remained unaffected, in contrast to other research findings. Although previous studies have reported a decline in the incidence of bacterial pneumonia [14], this study has not exhibited any decrease in its occurrence during the COVID-19 pandemic. Despite rigorous post-COVID-19 pandemic infection control measures, the persistent incidence of bacterial pneumonia indicates that the most significant risk factor for bacterial CAP in HM patients is their immune status [18].

As mentioned earlier, the prevalence of viral pneumonia decreased significantly during the COVID-19 pandemic. Bilateral pulmonary infiltration on chest X-ray also showed a significant decrease after the COVID-19 outbreak, which was hypothesised to be a result of a lower incidence of viral pneumonia [19]. Respiratory viruses usually cause mild upper respiratory tract infections; however, previous studies have reported that respiratory viruses are important pathogens of CAP in immunocompromised patients and elderly individuals [20,21]. As reported in other studies, the implementation of various preventive measures following the COVID-19 outbreak is likely to not only curtail the transmission of COVID-19 but also reduce the incidence of viral pneumonia caused by other respiratory viruses [2-4]. In this study, we observed an increase in the incidence of RSV during the winter of 2021, which is likely attributable to the RSV outbreak in South Korea due to the relaxation of domestic infection control measures in November 2021 [22].

In this study, a substantial number of patients (39.3%) presented with CAP of an undetermined aetiology. The prevalence of CAP of unknown aetiology among immunocompromised patients is relatively common [23,24]. Despite including only patients who underwent BRS to minimise the number of cases with an undetermined aetiology, the findings of this study still revealed this outcome. Given that the incidence of CAP of undetermined aetiology did not decrease during the COVID-19 pandemic, it is more likely that these cases were not caused by pathogens such as viruses or bacteria. Instead, it may be associated with the patients’ underlying disease or their immunocompromised status.

We investigated the risk factors for in-hospital mortality in HM patients with CAP. Consistent with previous studies [25,26], higher SOFA scores and lower platelet counts were found to be risk factors for in-hospital mortality in HM patients with CAP. In general, while the CURB-65 criteria or pneumonia severity index (PSI) are commonly used to predict the prognosis of patients with pneumonia, previous studies have shown that they are not effective in immunocompromised patients with cancer [27]. Thus, we opted to use the SOFA score instead. Thrombocytopenia is also known to reflect the severity and poor prognosis of infections [28,29]. Platelets play a role in inflammation and host defence mechanisms against microbial agents. Additionally, thrombocytopenia reflects bone marrow failure in patients with HM [30]. Therefore, thrombocytopenia could be a significant predictor of poor prognosis in patients with HM and CAP.

This study has several limitations. First, its retrospective design and single-centre implementation may introduce a selection bias, potentially affecting the significance of our findings. Nevertheless, we meticulously examined all admitted patients with CAP who underwent BRS in this single, large, 4-year cohort with consistent treatment protocols. The objective of our study was to assess the impact of the COVID-19 pandemic on the changing pattern of aetiology of CAP in patients with HM; thus, the setting of this study did not significantly deviate from that of a prospective observational study. Second, conducting a single-centre study may have had limitations in determining the overall aetiology of CAP in all patients with HM. However, this institution serves as the largest Asian centre for haematological disorders, performing over 500 bone marrow transplantations annually. Therefore, these data can be considered reasonably representative. Third, patients’ COVID-19 histories were not investigated. Previous studies reported that COVID-19 is a risk factor for bacterial or fungal co-infections and can play a significant role in patient prognosis [31,32]. However, due to the retrospective nature of the study, it was not possible to conduct an investigation. Additional studies are needed to evaluate the impact of a COVID-19 history on the prognosis of patients with CAP and HM.

## Conclusions

In the context of pandemic eras such as COVID-19, understanding the changes in CAP aetiology among immunocompromised patients, including those with HM, is crucial. The CAP incidence in patients with HM did not decrease after COVID-19, unlike that in the general population. A notable shift in the aetiology of CAP emerged among patients with HM following the COVID-19 pandemic, with a significant reduction in viral pneumonia but persistence of bacterial and fungal pneumonia. Further research is needed to evaluate the effects of COVID-19 on the prognosis of patients with HM and CAP.

## Data Availability

All data produced in the present work are contained in the manuscript.

## Acknowledgements

None

---

**Table S1.**
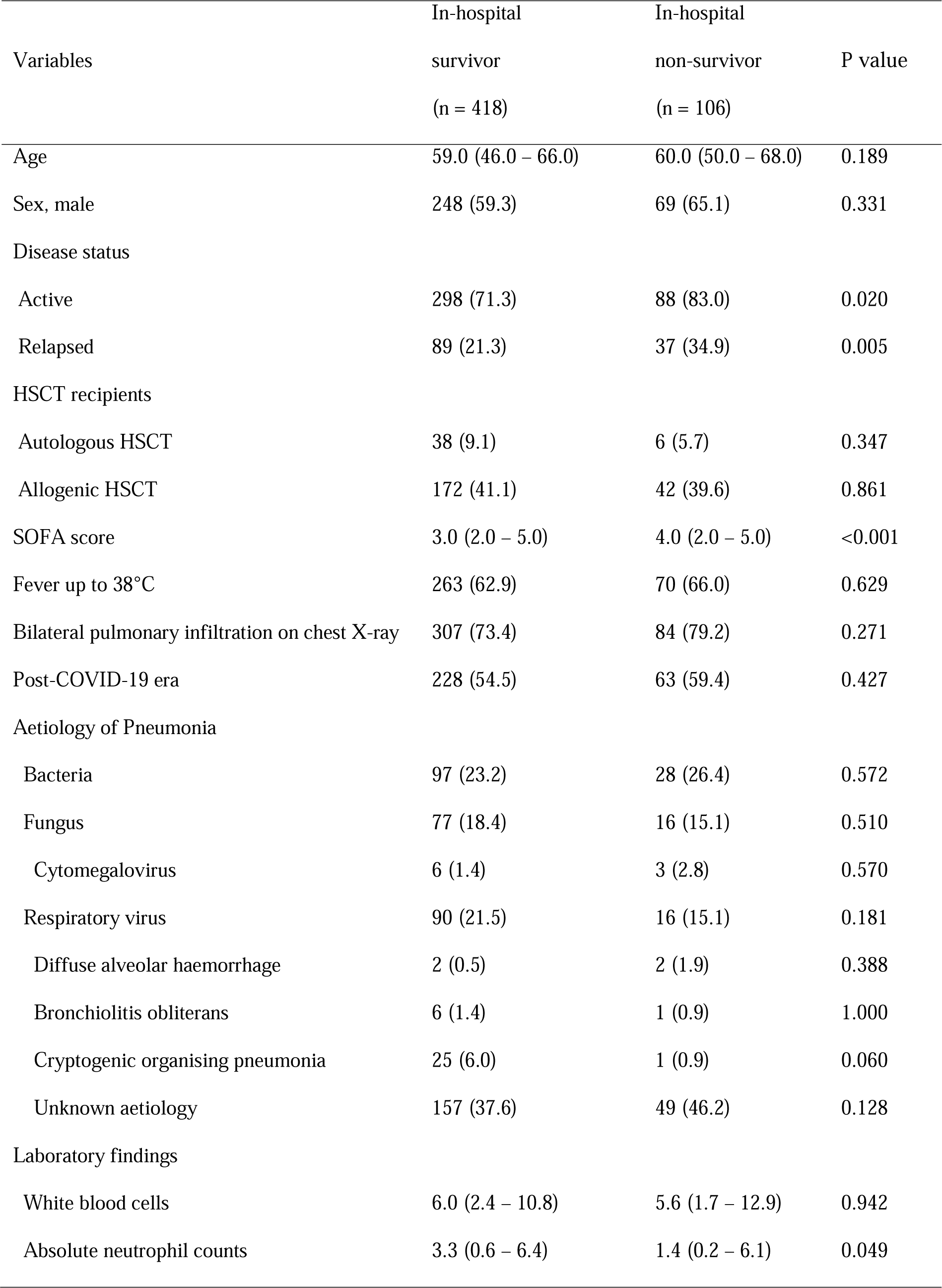
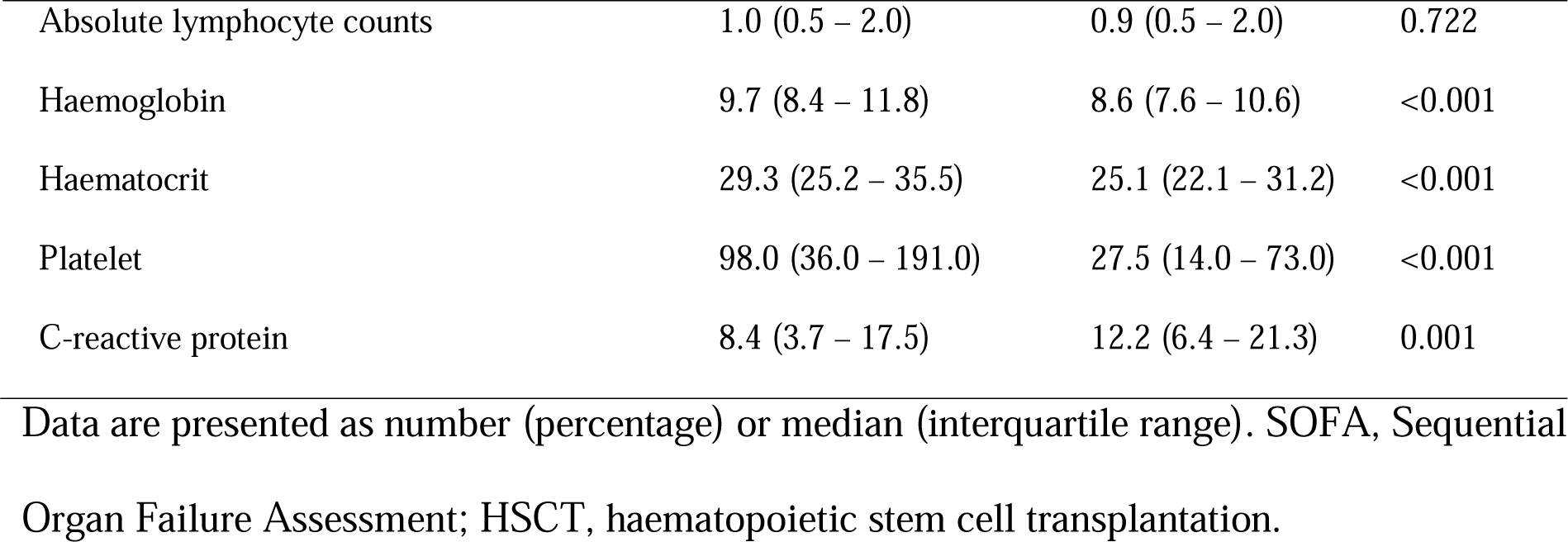
Clinical factors affecting in-hospital mortality.

